# Effectiveness of 2024/25 KP.2 vaccine against outpatient COVID-19 in Canada

**DOI:** 10.1101/2025.09.05.25335203

**Authors:** Lea Separovic, Suzana Sabaiduc, Yuping Zhan, Samantha E Kaweski, Sara Carazo, Romy Olsha, Richard G Mather, Christine Lacroix, Maan Hasso, Inès Levade, Isabelle Meunier, Agatha N Jassem, Katie Dover, Ruimin Gao, Nathalie Bastien, Danuta M Skowronski

## Abstract

Between November 2024 and April 2025, the Canadian Sentinel Practitioner Surveillance Network assessed KP.2 vaccine effectiveness against outpatient COVID-19 by test-negative design. Vaccination protected best during the first two months, reducing risk by two-thirds among vaccinated versus unvaccinated individuals, declining by 3-4 months, and reducing the risk overall by half.

## Background

Prior to the 2024/25 respiratory season, SARS-CoV-2 circulation in the United States (US) and Canada peaked between August-September 2024, followed by lower-level circulation compared to previous seasons [1]. Whereas 2024/25 vaccine strains elsewhere (e.g., Europe, US [2,3]) included JN.1 or KP.2, in Canada, a universal fall vaccination campaign offered publicly-funded mRNA vaccines containing KP.2 only. Ultimately, circulating viruses were mostly comprised of other JN.1 descendant or recombinant strains such as KP.3, XEC, and their sub-lineages [4]. Few estimates of 2024/25 COVID-19 vaccine effectiveness (VE) are available to date, and most have only reported interim season findings. Here, the Canadian Sentinel Practitioner Surveillance Network (SPSN) estimates end-of-season 2024/25 KP.2 VE against outpatient COVID-19 including genetic characterization of contributing case viruses and exploration of waning effects.

## Methods

We estimated COVID-19 VE by test-negative design among patients presenting to community-based sentinel practitioners in the provinces of British Columbia (BC), Ontario, and Quebec within 7 days of onset of acute respiratory illness (ARI; new or worsening cough potentially due to infection). Respiratory specimens were tested by accredited provincial laboratories using molecular detection assays. Fall KP.2 vaccination began late-September to mid-October, 2024. We include specimens collected between epi-weeks 44-18 (27 October 2024 to 03 May 2025). Ethical review waivers were provided in participating provinces.

In primary analyses, vaccine status was based upon information from provincial immunization registries (PIR), as previously [5]. In supplementary analyses, we compare VE based on self-report. We exclude children <12 years (due to more complex dosing recommendations), influenza-positive controls (to address correlated vaccination behaviours) and those vaccinated <2 weeks before onset or with unknown vaccine status or timing [6]. Logistic regression estimated odds ratios (OR), comparing likelihood of vaccination among test-positive cases versus test-negative controls, adjusting for age group, province, and calendar time. VE was derived as (1-OR)*100%. Variant contribution was based upon whole genome sequencing of case viruses (GISAID Epi_Set_ID: EPI_SET_250903wr), assigning SARS-CoV-2 lineages using Pango nomenclature (**Supplementary Table 1**). VE by variant and time since vaccination (TSV) were assessed as feasible.

## Results

Participant characteristics are detailed in **Supplementary Table 2**. Cases were slightly older than controls (median ages 49 vs 46). The proportion vaccinated among controls (19%) was comparable to most recently available 2023/24 fall campaign vaccine coverage in Canada (19% among those ≥5 years)[7].

Percent SARS-CoV-2 positivity among SPSN specimens remained low and stable at ~10% between epi-weeks 44-7, declining to ≤5% most weeks thereafter (**Supplementary Figure 1**). We successfully sequenced 312/435 (72%) case viruses, with consistent mix of multiple JN.1 sub-lineages and recombinants, with no single dominant escape variant emerging (**Supplementary Figure 2**). Over one third were XEC (35%; 110/312) or KP.3.1.1 (38%; 118/312, including descendant MC). XEC contribution successively increased between epi-weeks 44-46 (21%; 7/34), 47-49 (32%; 12/37), and 50-52 (49%; 21/43), remaining ≤50% thereafter. KP.3.1.1 contribution decreased during the same respective periods (71% (24/34), 54% (20/37), and 37% (16/43)), with equal proportions ancestral KP.3.1.1 (52%; 31/60) and descendant MC (48%; 29/60) across epi-weeks 44-52, but with lower KP.3.1.1 (18%; 9/51) than MC (80%; 41/51) between epi-weeks 1-9. The emerging LP.8.1 variant comprised just 12% (37/312) of sequenced viruses overall, reaching 33% (9/27) during epi-weeks 10-18 but with low case detection overall.

At median time post-vaccination of 10-11 weeks, VE against outpatient COVID-19 was 54%, somewhat lower when restricted to ≥65-year-olds (40%)(**Figure**). Overall, VE decreased from 68% at 2-7 weeks to 27% at ≥12 weeks (median 17, IQR 14-20, range 12-28 weeks), notably negligible at ≥16 weeks (median 19, IQR 17-22 weeks) post-vaccination. With comparable TSV (median 10-11 weeks), variant-specific XEC versus non-XEC estimates were similar (45% versus 48%, respectively). In sensitivity analyses (**Supplementary Tables 3-4**), VE changed minimally (absolute difference) when including flu-positive controls (−3% to −6%), excluding participants vaccinated ≤6 months before fall vaccine campaign (+5% to +9%), or adjusting for sex and comorbidity (−1%), slightly more for self-report versus PIR-based VE (−12% to +3%).

**Figure.**
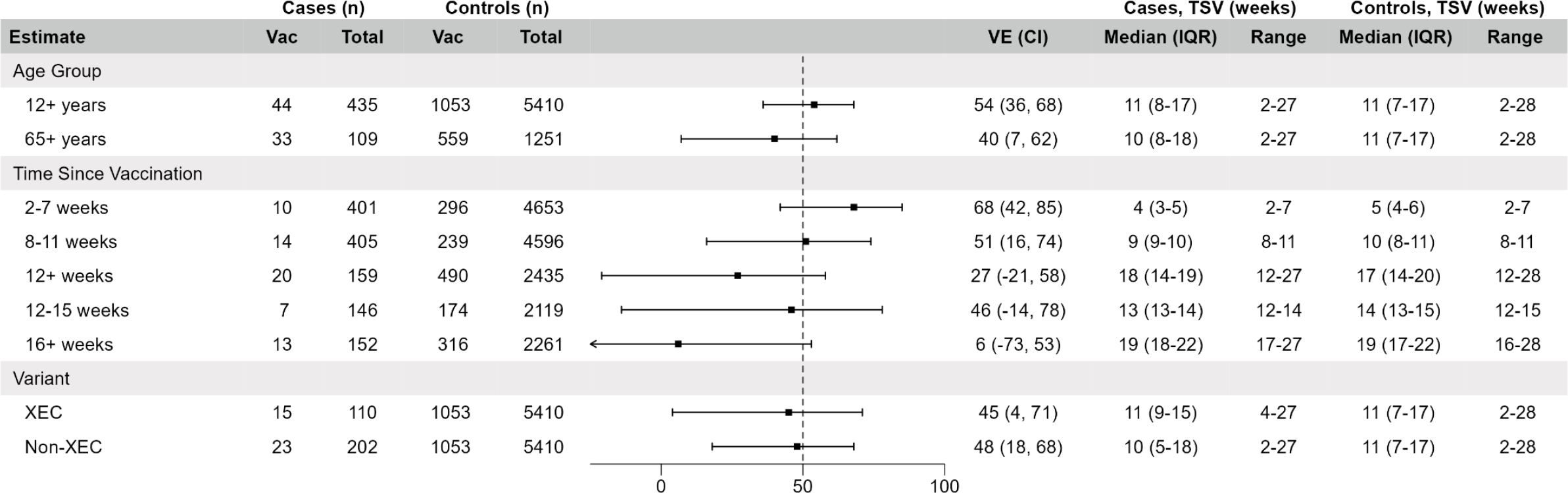
Vaccine effectiveness against acute respiratory illness due to SARS-CoV-2, Canadian Sentinel Practitioner Surveillance Network (SPSN), 27 October 2024 to 03 May 2025 (epi-weeks 44-18) (n=5845) Abbreviations: CI, confidence interval; IQR, interquartile range; TSV, time since vaccination; Vac, vaccinated; VE, vaccine effectiveness. Odds ratios (ORs) were estimated with logistic regression adjusting for province (BC, Ontario, Quebec), calendar time (bi-weekly epiweeks), and age group (12-49, 50-64, and 65+ years). VE derived as (1-OR)*100%. All estimates are for those ≥12 years unless otherwise specified. Vaccinated participants are those who received Fall 2024/25 vaccine ≥2 weeks prior to ARI onset. No participants in VE analyses received a Spring 2025 booster dose prior to ARI onset. Time since vaccination refers to interval between receipt of last COVID-19 vaccine and ARI onset among vaccinated participants. Analyses span epi-weeks 44 to 18 but due to few vaccinated participants reaching 12+ weeks since vaccination during the first half of the season (n=28 in epi-weeks 44 to 4 versus n=510 in epi-weeks 5 to 18), we restricted the analysis of VE at 12+, 12-15, and 16+ weeks post-vaccination to epi-weeks 5 to 18. Minimal difference in VE (≤1% absolute) was observed when using Firth’s logistic regression to address small sample size. For details on virological characterization, see Supplementary Table 1.

## Discussion

The community-based Canadian SPSN reports the 2024/25 KP.2 mRNA vaccine halved the risk of COVID-19 overall among ≥12-year-olds requiring outpatient medical visit for ARI. These findings of substantial risk reduction are especially meaningful given that by 5 years post-pandemic the immunological landscape has dramatically changed, with most of the residual COVID-19 healthcare burden now likely concentrated in outpatient settings [8]. As previously [9], we show protection was greatest within two months post-vaccination, reducing the risk by about two-thirds, but rapidly declined to negligible by 3-4 months post-vaccination.

With nearly three-quarters of case viruses sequenced, we directly contextualize our VE findings within the mix of contributing variants. XEC and KP.3.1.1 variants were most common, each comprising about one-third. In addition to the FLiRT/FLiQE substitutions T346R and Q493E relative to vaccine, both variants separately acquired N-terminal domain substitutions and glycans of potential relevance to immune evasion [10,11]. While both JN.1 and KP.2 vaccines elicit robust neutralizing antibody titres against all JN.1 sublineages (slightly higher for KP.2 vaccines), modest reductions against KP.3 and more so against XEC/KP.3.1.1 have been reported [12–15]. Absent serological thresholds for protection, the extent to which immunogenicity differences may explain suboptimal (albeit substantial) VE, even shortly post-vaccination, remains unclear. Ideally, analysis of waning protection should simultaneously account for concomitant changes in variant contribution [9]. For the current work this includes early but brief predominance of KP.3.1.1 and stable but slightly increasing XEC contribution across the analysis period. Although sample size precluded finer TSV stratification, similar VE for XEC (45%) and non-XEC (48%) variants at comparable median TSV suggests the interpretation of waning is not driven by related shifts in variant contribution. Although XEC increased across the season, it never comprised >50% of variants, further reinforcing it was not a strong contender for immune escape through the 2024/25 season.

In 2024/25, findings from England, Denmark, and the US instead report sustained protection with stable VE after ≥20, ≥13, and a range of ~8-17 weeks post-vaccination, respectively [16–18]. Contextual differences, including the relative mix of vaccine products, participant ages, weighting within TSV strata, and contribution and timing of variant circulation, must be considered when comparing VE across jurisdictions. For instance, US estimates may reflect a mix of mRNA and protein subunit vaccines (although in unknown proportions), with waning more evident in younger adults (18-64 years) than overall (≥18 years)[16]. Also, with median ~11 weeks and IQR of just 10-13 weeks post-vaccination, the longest TSV stratum within their September-January analysis period may be weighted by participants with TSV still within the period for which we also observed substantial protection. In our study, most participants with ARI onset ≥12 weeks post-vaccination presented between February-April. Better understanding of the duration of protection is critical to optimal immunization program timing, including staggered target groups and necessity of same-season (e.g., spring) boosters.

Most studies of 2024/25 COVID-19 VE have focused on older adults. Estimated VE among ≥65-year-olds in our study (40%) is comparable to older adult estimates from other test-negative design studies, including against outpatient illness in the US VISION network (~35%)[16] and hospitalization in England (~35-45%)[17], albeit lower than elsewhere, with VE exceeding 65% against outpatient illness in Europe [19] and hospitalization in a Danish cohort study [18]. As mentioned, other differences must be considered when comparing VE estimates, including methodological. For instance, the higher KP.2 VE reported in the US Veteran Affairs healthcare system (58%) was not adjusted for calendar time, despite indication of confounding on that basis, and reflects short-term protection (median ~4 weeks post-vaccination) from September-November [20]. Vaccine status ascertainment (e.g., self-report, registry) and vaccine strain also varied elsewhere. In our study we observed VE under-estimation (up to 12% absolute) with self-report, similar to findings elsewhere, reflecting misclassification bias with effects exacerbated by low vaccine coverage [21,22]. Epidemiologically, the impact of vaccine strain remains uncertain with wide range of strain-specific estimates (e.g., 40-70% JN.1 VE against inpatient illness [16,17] and 40-60% KP.2 VE against outpatient illness[20]), but none directly comparing relative JN.1 versus KP.2 VE.

The main limitation of the current study is the wide confidence intervals, with low-level SARS-CoV-2 activity and low vaccine coverage limiting statistical power and precluding further stratified analyses. We uniquely provide detailed genetic characterization of contributing case viruses to help disentangle the effects of waning from differential variant-specific contribution across the season; however, we cannot draw definitive conclusions. VE estimates for the 2024/25 season may also require consideration of the shortly preceding summer peak in SARS-CoV-2 activity, and potential impact in lowering VE estimates.

Overall, end-of-season findings from the Canadian SPSN add to the limited body of literature otherwise available for the post-pandemic performance of seasonal COVID-19 vaccination. We show COVID-19 vaccination continues to provide added protection within a SARS-CoV-2-experienced population, with the 2024/25 KP.2 vaccine reducing the risk of outpatient ARI due to SARS-CoV-2 by about half overall and by two thirds within two months post-vaccination. As previously, however, we note protection waned considerably by four months post-vaccination with implications for pursuit of improved vaccine technologies and booster dose recommendations.

## Data Availability

Data for SPSN SARS-CoV-2 viruses meeting provincial and/or national criteria for upload and their submitting and contributing laboratories can be found on GISAID using the Epi_Set_ID: EPI_SET_250903wr.

