## Supplementary Materials for "Effectiveness of 2024/25 KP.2 vaccine against outpatient COVID-19 in Canada"

**Supplementary Table 1.** Lineage distribution of sequenced SARS-CoV-2 case viruses included in vaccine effectiveness analysis by province, Canadian Sentinel Practitioner Surveillance Network (SPSN), 27 October 2024 to 03 May 2025 (weeks 44-18)

| | | BC N = 49 | Ontario N = 277 | Québec N = 109 | TOTAL N = 435 |
|---|---|---|---|---|---|
| **Case viruses successfully sequenced, n (% n/N)** | | **42 (86%)** | **185 (67%)** | **85 (78%)** | **312 (72%)** |
| **Parental lineage[1, 2]** | **Sub-lineages detected among SPSN case viruses[2, 3]** | | | | |
| JN.1 | KS.1.1, LB.1.3.1, LF.7.*, MV.1, NC.1.2.2, NL.*, PC.*, PY.2 | 2 | 14 | 8 | 24 (8%) |
| LP.8.1 | NY.*, PD.1, PR.2 | 6 | 23 | 8 | 37 (12%) |
| KP.2 | NM.2 | | | 1 | 1 (0%) |
| KP.3 | NP.1, PG.6 | 2 | 5 | 1 | 8 (3%) |
| KP.3.1.1 | | 3 | 26 | 12 | 41 (13%) |
| MC | PA.1.4, PJ.1 | 7 | 43 | 27 | 77 (25%) |
| XEC | | 18 | 68 | 24 | 110 (35%) |
| Other | XEK.*, XEL.3, XEU, XFG, XFJ.1, NB.1.8.1[4], PQ.1.1[5] | 4 | 6 | 4 | 14 (4%) |

Abbreviations: BC, British Columbia; SPSN, Canadian Sentinel Practitioner Surveillance Network.
Whole genome sequencing of SPSN SARS-CoV-2 case viruses followed routine provincial or national laboratory protocols [1–6] and lineages assigned based on contemporary Pango nomenclature [7,8]. Data for SPSN SARS-CoV-2 viruses meeting provincial and/or national criteria for upload and their submitting and contributing laboratories can be found on GISAID using the Epi_Set_ID: EPI_SET_250903wr (https://doi.org/10.55876/gis8.250903wr) [9].

[1] All lineages include their descendants unless otherwise noted
[2] Lineages determined using pangolin version 4.3.1 [7]
[3] Lineages suffixed with an asterisk also include their descendant lineages, unless otherwise noted
[4] Descendant of XDV viruses; two detected in weeks 17-18
[5] Descendant of NB.1.8.1 viruses; one detected in week 14

**Supplementary Figure 1.** Epidemic curve of SARS-CoV-2 cases and controls, Canadian Sentinel Practitioner Surveillance Network (SPSN), 27 October 2024 to 03 May 2025 (weeks 44-18)

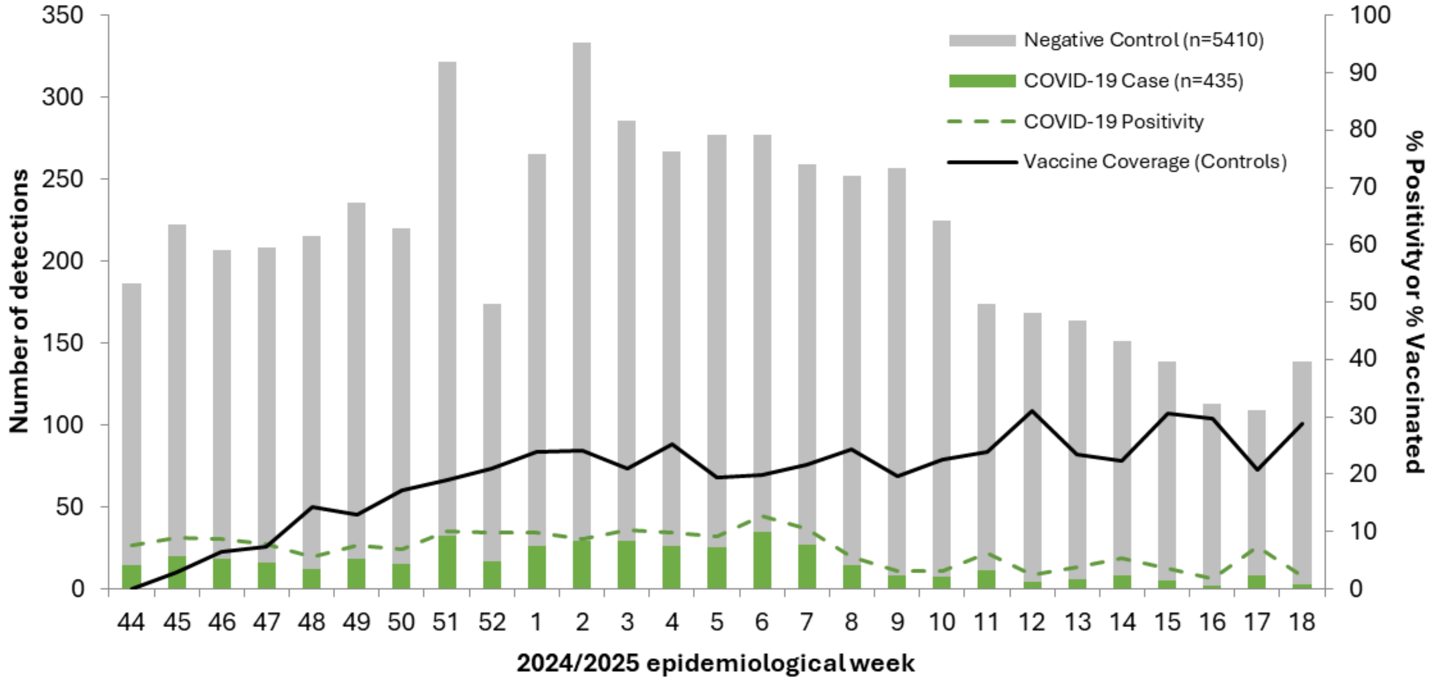

Displayed proportion vaccinated is based on those vaccinated ≥ 2 weeks before illness onset date.

**Supplementary Table 2.** Participant profile, COVID-19 vaccine effectiveness analysis, Canadian Sentinel Practitioner Surveillance Network (SPSN), 27 October 2024 to 03 May 2025 (weeks 44-18)

| Characteristics | All ARI participants (column %) | | | | | | | Proportion COVID-19 vaccinated[a] (row %) | | | | | | |
|---|---|---|---|---|---|---|---|---|---|---|---|---|---|---|
| | Overall | | COVID-19 cases | | COVID-19 controls | | p value[b] | Overall | | p value[c] | COVID-19 cases | | COVID-19 controls | |
| | n | % | n | % | n | % | | n | % | | n | % | n | % |
| N (row %) | 5845 | 100 | 435 | 7 | 5410 | 93 | NA | 1097 | 19 | NA | 44 | 10 | 1053 | 19 |
| **Age group (years)** | | | | | | | | | | | | | | |
| 12-49 | 3207 | 55 | 223 | 51 | 2984 | 55 | | 264 | 8 | | 7 | 3 | 257 | 9 |
| 50-64 | 1278 | 22 | 103 | 24 | 1175 | 22 | 0.292 | 241 | 19 | <0.001 | 4 | 4 | 237 | 20 |
| ≥ 65 | 1360 | 23 | 109 | 25 | 1251 | 23 | | 592 | 44 | | 33 | 30 | 559 | 45 |
| Median (IQR) | 47 (33-63) | | 49 (36-65) | | 46 (33-63) | | 0.002 | 66 (64-79) | | <0.001 | 72 (64-79) | | 66 (50-75) | |
| **Sex** | | | | | | | | | | | | | | |
| Female | 3744 | 64 | 278 | 64 | 3466 | 64 | 0.889 | 732 | 20 | 0.046 | 28 | 10 | 704 | 20 |
| Male | 2073 | 35 | 156 | 36 | 1917 | 35 | | 361 | 17 | | 15 | 10 | 346 | 18 |
| Unknown | 28 | 0 | 1 | 0 | 27 | 0 | NA | 4 | 14 | NA | 1 | 100 | 3 | 11 |
| **Comorbidity[d]** | | | | | | | | | | | | | | |
| No | 3839 | 66 | 288 | 66 | 3551 | 66 | 0.5 | 520 | 14 | <0.001 | 25 | 9 | 495 | 14 |
| Yes | 1580 | 27 | 127 | 29 | 1453 | 27 | | 457 | 29 | | 18 | 14 | 439 | 30 |
| Unknown | 426 | 7 | 20 | 5 | 406 | 8 | NA | 120 | 28 | NA | 1 | 5 | 119 | 29 |
| **Province** | | | | | | | | | | | | | | |
| British Columbia | 1452 | 25 | 49 | 11 | 1403 | 26 | | 354 | 24 | | 11 | 22 | 343 | 24 |
| Ontario | 2671 | 46 | 277 | 64 | 2394 | 44 | <0.001 | 423 | 16 | <0.001 | 28 | 10 | 395 | 16 |
| Quebec | 1722 | 29 | 109 | 25 | 1613 | 30 | | 320 | 19 | | 5 | 5 | 315 | 20 |
| **Epi-week of specimen collection, 2024/25[e]** | | | | | | | | | | | | | | |
| 44-45 | 408 | 7 | 34 | 8 | 374 | 7 | | 6 | 1 | | 0 | 0 | 6 | 2 |
| 46-47 | 415 | 7 | 34 | 8 | 381 | 7 | | 28 | 7 | | 2 | 6 | 26 | 7 |
| 48-49 | 451 | 8 | 30 | 7 | 421 | 8 | | 60 | 13 | | 3 | 10 | 57 | 14 |
| 50-51 | 542 | 9 | 47 | 11 | 495 | 9 | | 92 | 17 | | 2 | 4 | 90 | 18 |
| 52-1 | 439 | 8 | 43 | 10 | 396 | 7 | | 95 | 22 | | 5 | 12 | 90 | 23 |
| 2-3 | 619 | 11 | 58 | 13 | 561 | 10 | | 132 | 21 | | 5 | 9 | 127 | 23 |
| 4-5 | 544 | 9 | 51 | 12 | 493 | 9 | <0.001 | 113 | 21 | <0.001 | 3 | 6 | 110 | 22 |
| 6-7 | 536 | 9 | 62 | 14 | 474 | 9 | | 107 | 20 | | 9 | 15 | 98 | 21 |
| 8-9 | 509 | 9 | 22 | 5 | 487 | 9 | | 112 | 22 | | 5 | 23 | 107 | 22 |
| 10-11 | 399 | 7 | 18 | 4 | 381 | 7 | | 89 | 22 | | 1 | 6 | 88 | 23 |
| 12-13 | 332 | 6 | 10 | 2 | 322 | 6 | | 90 | 27 | | 2 | 20 | 88 | 27 |
| 14-15 | 290 | 5 | 13 | 3 | 277 | 5 | | 75 | 26 | | 2 | 15 | 73 | 26 |
| 16-18 | 361 | 6 | 13 | 3 | 348 | 6 | | 98 | 27 | | 5 | 38 | 93 | 27 |
| **Received COVID-19 vaccine from April to September in 2024[f]** | | | | | | | | | | | | | | |
| Yes | 247 | 4 | 17 | 4 | 230 | 4 | 0.765 | 168 | 68 | <0.001 | 11 | 65 | 157 | 68 |
| No | 5598 | 96 | 418 | 96 | 5180 | 96 | | 929 | 18 | | 33 | 9 | 896 | 19 |

ARI: Acute respiratory illness; IQR: interquartile range; NA: not applicable. Unless otherwise specified, values displayed in the columns represent the number of specimens per category and percentages are relative to the total.

[a] Vaccination status refers to vaccination as part of Fall 2024/25 vaccine campaigns, based on provincial immunization registry. Participants vaccinated < 2 weeks before onset of symptoms or with unknown vaccination status or timing were excluded. No participants in VE analyses received Spring 2025 booster vaccine prior to symptom onset.

[b] p values for comparison between cases and controls were derived by two-way chi-squared test or Wilcoxon rank-sum test.

[c] p values for comparison between vaccinated and not vaccinated were derived by two-way chi-squared test or Wilcoxon rank-sum test. The number not vaccinated can be derived by subtracting the number vaccinated from the total ARI participants, by row.

[d] Includes chronic comorbidities that place individuals at higher risk of serious complications from influenza as defined by Canada's National Advisory Committee on Immunization [10].

[e] Missing specimen collection dates were imputed as the date the specimen was received and processed at the laboratory minus 2 days.

[f] Participants vaccinated between 1 April 2024 to 30 September 2024. Earliest start date for Fall 2024/25 vaccine campaigns in SPSN provinces was 30 September 2024 for high-risk groups in Ontario; however, all vaccinated SPSN participants received Fall dose on or after 1 Oct 2024.

**Supplementary Figure 2**. Lineage distribution of sequenced SARS-CoV-2 viruses, Canadian Sentinel Practitioner Surveillance Network (SPSN), 27 October 2024 to 03 May 2025 (weeks 44-18).

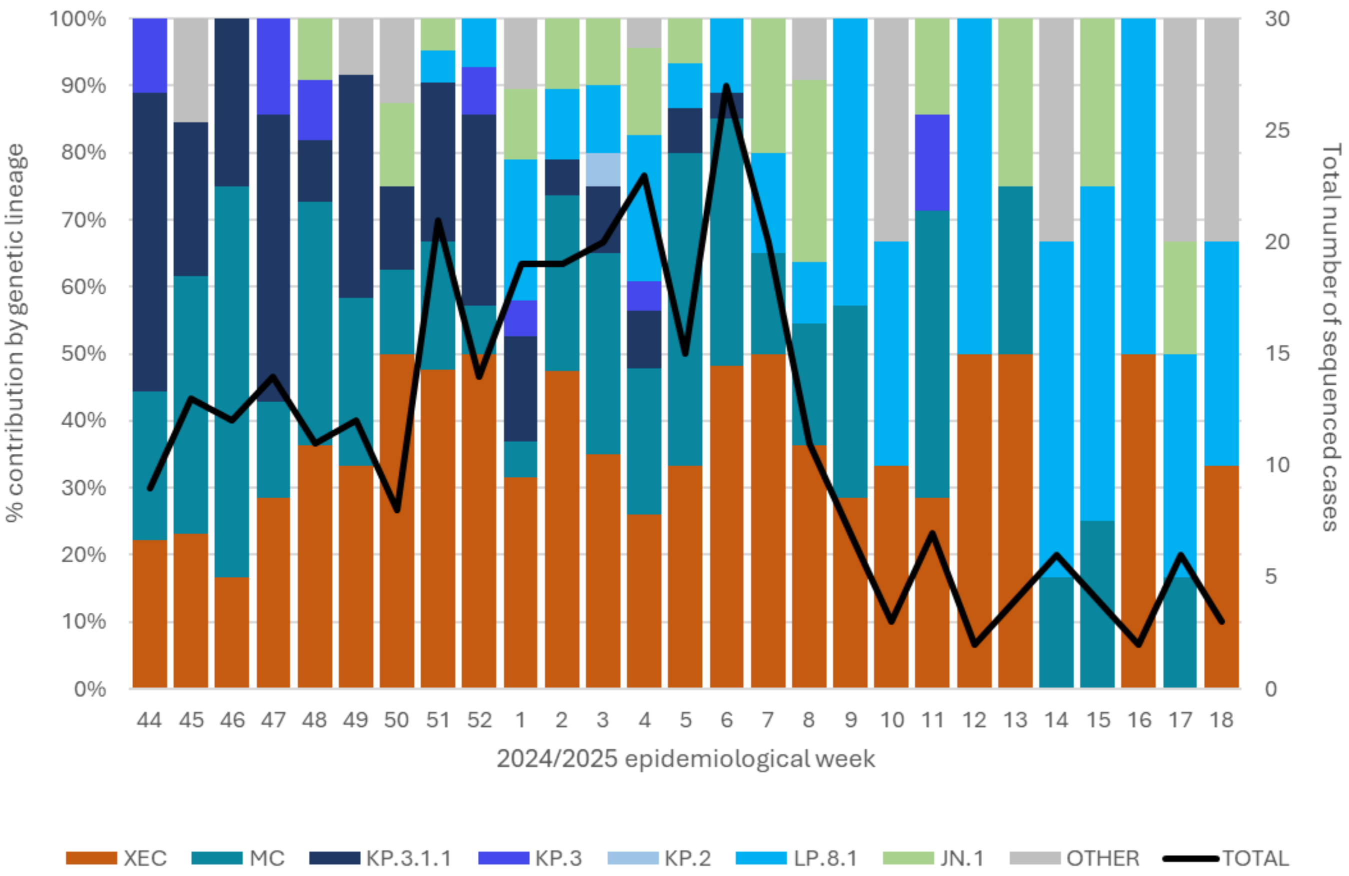

Displayed is the genomic distribution of SARS-CoV-2 case viruses (n=312) contributing to SPSN COVID-19 VE analysis. All viral lineages displayed are descendants or recombinants of JN.1 lineages, except for recombinants denoted as 'Other'. See **Supplementary Table 1** for more details.

**Supplementary Table 3.** Association between vaccination and acute respiratory illness due to SARS-CoV-2, sensitivity analyses, Canadian Sentinel Practitioner Surveillance Network (SPSN), 27 October 2024 to 03 May 2025 (weeks 44-18)

| | Total | Cases | | Controls | | Unadjusted OR[a] | | Adjusted OR[a,b] | | Adjusted VE[a,b] | |
|---|---|---|---|---|---|---|---|---|---|---|---|
| | N | n vac[c]/N | % | n vac[c]/N | % | OR | 95% CI | OR | 95% CI | VE | 95% CI |
| **Primary analysis** | | | | | | | | | | | |
| ≥ 12 years | 5845 | 44/435 | 10 | 1053/5410 | 19 | 0.47 | (0.33, 0.63) | 0.46 | (0.32, 0.64) | 54 | (36, 68) |
| ≥ 65 years | 1360 | 33/109 | 30 | 559/1251 | 45 | 0.54 | (0.35, 0.81) | 0.60 | (0.38, 0.93) | 40 | (7, 62) |
| **Adjusted for sex and comorbidity[d]** | | | | | | | | | | | |
| ≥ 12 years | 5394 | 42/414 | 10 | 931/4980 | 19 | 0.49 | (0.35, 0.67) | 0.47 | (0.33, 0.66) | 53 | (34, 67) |
| ≥ 65 years | 1225 | 32/107 | 30 | 481/1118 | 43 | 0.57 | (0.36, 0.86) | 0.61 | (0.39, 0.95) | 39 | (5, 61) |
| **Excluding those receiving a COVID-19 vaccine between April to September 2024** | | | | | | | | | | | |
| ≥ 12 years | 5598 | 33/418 | 8 | 896/5180 | 17 | 0.41 | (0.28, 0.58) | 0.41 | (0.27, 0.59) | 59 | (41, 73) |
| ≥ 65 years | 1169 | 22/95 | 23 | 430/1074 | 40 | 0.45 | (0.27, 0.73) | 0.51 | (0.30, 0.84) | 49 | (16, 70) |
| **Including influenza-positive SARS-CoV-2 controls** | | | | | | | | | | | |
| ≥ 12 years | 8101 | 44/435 | 10 | 1327/7666 | 17 | 0.54 | (0.39, 0.73) | 0.52 | (0.36, 0.72) | 48 | (28, 64) |
| ≥ 65 years | 1667 | 33/109 | 30 | 688/1558 | 44 | 0.55 | (0.36, 0.83) | 0.63 | (0.41, 0.98) | 37 | (2, 59) |

Abbreviations: CI, confidence interval; OR, odds ratio; vac, vaccinated; VE, vaccine effectiveness.

[a] ORs compared test positivity between vaccinated and unvaccinated participants with logistic regression. VE derived as (1 – OR) x 100.

[b] Adjusted for age group (12-49, 50-64, ≥65 years), province (BC, Ontario, Quebec), and calendar time (bi-weekly epi-weeks).

[c] Vaccination status based on provincial immunization registry. Participants vaccinated < 2 weeks before onset of symptoms or with unknown vaccination status or timing were excluded.

[d] Participants with unknown sex or comorbidity status were excluded.

**Supplementary Table 4.** Association between vaccination and acute respiratory illness due to SARS-CoV-2, vaccine status per provincial immunization registry or self-report, Canadian Sentinel Practitioner Surveillance Network (SPSN), 27 October 2024 to 19 April 2025 (weeks 44-16)

| | Total | Cases | | Controls | | Unadjusted OR[a] | | Adjusted OR[a,b] | | Adjusted VE[a,b] | |
|---|---|---|---|---|---|---|---|---|---|---|---|
| | N | n vac[c]/N | % | n vac[c]/N | % | OR | 95% CI | OR | 95% CI | VE | 95% CI |
| **Vaccination status per PIR[d]** | | | | | | | | | | | |
| ≥ 12 years | 5597 | 41/424 | 10 | 993/5173 | 19 | 0.45 | (0.32, 0.62) | 0.44 | (0.31, 0.62) | 56 | (38, 69) |
| ≥ 65 years | 1284 | 30/106 | 28 | 516/1178 | 44 | 0.51 | (0.32, 0.78) | 0.56 | (0.35, 0.87) | 44 | (13, 65) |
| **Vaccination status per self-report[d]** | | | | | | | | | | | |
| ≥ 12 years | 4995 | 55/391 | 14 | 1034/4604 | 22 | 0.57 | (0.42, 0.75) | 0.56 | (0.41, 0.77) | 44 | (23, 59) |
| ≥ 65 years | 1130 | 30/100 | 30 | 473/1030 | 46 | 0.50 | (0.32, 0.78) | 0.53 | (0.33, 0.83) | 47 | (17, 67) |

Abbreviations: CI, confidence interval; OR, odds ratio; PIR, provincial immunization registry; vac, vaccinated; VE, vaccine effectiveness.

[a] ORs compared test positivity between vaccinated and unvaccinated participants with logistic regression. VE derived as (1 – OR) x 100.

[b] Adjusted for age group (12-49, 50-64, ≥65 years), province (BC, Ontario, Quebec), and calendar time (bi-weekly epi-weeks).

[c] Vaccination status based on PIR or self-report as indicated. Participants vaccinated < 2 weeks before onset of symptoms or with unknown vaccination status or timing were excluded.

[d] Analyses restricted to epi-weeks 44-16 in order to exclude participants with most recent vaccine receipt potentially during Spring 2025 booster campaign, with rollout beginning in early April in SPSN provinces, given exact vaccination date unavailable for self-reported vaccination status.

### References, Supplementary Material
